# Qing Fei Pai Du Tang, a Chinese multi-herbal medicine formulated against COVID-19, elevates the plasma levels of IL-1β, IL-18, TNF-α, and IL-8

**DOI:** 10.1101/2020.07.13.20146175

**Authors:** Yasunari Kageyama, Koichi Aida, Kimihiko Kawauchi, Masafumi Morimoto, Tomoka Ebisui, Tetsu Akiyama, Tsutomu Nakamura

## Abstract

There are currently no specific vaccine or drugs proven to be effective against COVID-19. Traditional Chinese herbal medicine has been integrated into the official therapeutic protocol against COVID-19 in China. Qing Fei Pai Du Tang (QFPDT) is a Chinese multi-herbal formula newly developed and specifically optimized for the treatment of COVID-19. Therapeutic administration of QFPDT resulted in an improved cure rate in mild to critically-ill patients. However, the immunological mechanism for the efficacy of QFPDT has been poorly understood. Furthermore, the feasibility of prophylactic use in uninfected individuals remain unconfirmed. We thus examined whether the administration of QFPDT at a dose lower than recommended for therapeutic use alters hematological and/or immunological measures in healthy individuals. We found that QFPDT elevates the plasma levels of IL-1β, IL-18, TNF-α, and IL-8, which are key mediators of acute inflammatory responses to ssRNA viruses. No apparent adverse effects were observed during the trial. Our finding suggests that the pharmacological action of QFPDT is associated with the upregulation of a subset of proinflammatory cytokines despite its clinical benefits for COVID-19 patients. We should therefore be careful in its prophylactic use in uninfected individuals until we have a better understanding of the immunopharmacological action of QFPDT through further clinical studies with larger cohorts.

## Introduction

Currently effective vaccines or specific therapeutic agents against COVID-19 remain unavailable. The majority of patients with COVID-19 in China have been treated with a combination of traditional Chinese and modern Western medicine [1–3]. The use of several Chinese herbal formulas is encouraged for the treatment of COVID-19 in the latest version of the Diagnosis and Treatment Protocol for Novel Coronavirus Pneumonia (Trial Version 7) released by the National Health Commission of China. One of them is “Qing Fei Pai Du Tang” (QFPDT, the Chinese word for “lung cleansing and detoxifying decoction”), which is newly formulated and specifically optimized for the symptoms of COVID-19. QFPDT has shown a satisfactory therapeutic efficacy when administered to thousands of mild to critical cases [1–3]. However, the underlying pharmacological mechanism and the feasibility of prophylactic application to uninfected individuals remain unknown. We thus examined whether QFPDT administration at a dose lower than recommended for therapeutic use affects hematological and immunological measures in healthy individuals.

## Materials and Methods

### Statement of Ethics

This study was carried out in accordance with The Code of Ethics of the World Medical Association (Declaration of Helsinki). All procedures were approved by the Ethics Committees of the Takanawa Clinic (approval number: 2020-2). A signed informed consent form was obtained from each participant prior to inclusion in this study. All experimental procedures and data analyses were conducted by investigators who were blinded to the subjects’ clinical information using a de-identified dataset. This study has been registered on University Hospital Medical Information Network-Clinical Trials Registry (UMIN-CTR) under the trial number UMIN000040341.

### Subjects

This is an open-label, single-arm study to evaluate the feasibility of prophylactic use of QFPDT in healthy individuals and obtain a clue to the pharmacological action of QFPDT. A total of 18 healthy volunteers were enrolled (5 males, 13 females; ages 22–58 years; mean age [SD], 33.8 [10.7] years), all of whom tested negative for IgM and IgG antibodies to SARS-CoV-2. Individuals were excluded if they had current infectious, inflammatory, or immune-related diseases.

### Preparation and administration of QFPDT

Twenty-one kinds of traditional Chinese herbs for the QFPDT formula were purchased from Shanghai Ruisha Comlat Co., Ltd. (Shanghai, China). The herbs were mixed in accordance with the above-mentioned Chinese official guideline; however, we reduced the dose of each herb to 1/30 to explore the feasibility of prophylactic use and avoid possible adverse effects such as palpitation. The QFPDT decoction was prepared by boiling the mixed herbs in 600 ml of water for 1 h before the first administration (day 1), divided into six aliquots and stored at 4°C during the trial. The subjects were instructed to take an aliquot of the decoction orally 40 min after breakfast and dinner for 3 days (day 1–3). Peripheral blood samples were obtained 12 h before the first administration (day 0) and after the last administration (day 4).

### Hematological, biochemical and cytokine analyses

Hematological and biochemical tests were outsourced to SRL, Inc. (Tokyo, Japan). Plasma cytokines were quantified using V-PLEX Proinflammatory Panel 1 Human Kit (Meso Scale Diagnostics) and Human IL-18 ELISA Kit (Abcam). All collected data (n = 18) were subjected to the statistical analysis using a two-tailed paired *t*-test.

## Results

Blood urea nitrogen, mean corpuscular volume, and mean corpuscular hemoglobin concentration changed marginally within the normal ranges (Table 1), but there were no significant differences in other hematological and biochemical measures between day 0 and day 4. Notably, we found that the plasma levels of proinflammatory cytokines IL-1β, IL-18, TNF-α, and IL-8 in day 4 were significantly higher than those in day 0 (IL-1β: 0.173 vs. 0.558, *P* = 0.0198; IL-18: 119 vs. 269, *P* = 0.0207; TNF-α: 1.82 vs. 2.36, *P* = 0.00536; IL-8: 291 vs. 663, *P* = 0.00184; Table 1). These cytokines were increased in 14 (77.8%), 15 (83.3%), 15 (83.3%), and 17 (94.4%) out of 18 subjects, respectively (Fig. 1). We also found a slight increase in IL-6 (0.552 vs. 0.852), which is known to be closely associated with the severity and mortality of COVID-19 [4–6]; however, the difference was not statistically significant (*P* = 0.147). No apparent adverse effects were observed during the trial.

**Table 1.** Hematological and cytokine changes in QFPDT-administered healthy individuals.

| Parameters | Day 0 |  | Day 4 |  | Mean (Day 4)<br>– Mean (Day 0) | 95% CI | P-value |
| --- | --- | --- | --- | --- | --- | --- | --- |
|  | Mean | SD | Mean | SD |  |  |  |
| Complete blood count |  |  |  |  |  |  |  |
| Red blood cell count (x 10<sup>4</sup>/μl) | 452 | 44.4 | 449 | 45.3 | -3.06 | -11.7–5.57 | 0.465 |
| Hemoglobin (g/dl) | 13.5 | 1.78 | 13.5 | 1.79 | -0.0556 | -0.347–0.236 | 0.692 |
| Hematocrit (%) | 40.8 | 4.58 | 41.4 | 4.68 | 0.578 | -0.370–1.53 | 0.216 |
| MCV (fl) | 90.4 | 6.31 | 92.3 | 6.69 | 1.93 | **0.908–2.96** | **0.000971\*\*\*** |
| MCH (pg) | 29.9 | 2.60 | 30.0 | 2.72 | 0.106 | -0.0878–0.299 | 0.265 |
| MCHC (%) | 33.1 | 1.39 | 32.5 | 1.38 | -0.600 | **-0.946 to -0.254** | **0.00194\*\*** |
| White blood cell count (/μl) | 6160 | 1240 | 5890 | 1270 | -267 | -692–158 | 0.203 |
| Platelet count (x 10<sup>4</sup>/μl) | 25.1 | 3.50 | 24.5 | 3.74 | -0.511 | -1.40–0.373 | 0.239 |
| White blood cell differential |  |  |  |  |  |  |  |
| Neutrophils (%) | 59.2 | 6.86 | 57.9 | 7.83 | -1.29 | -4.04–1.46 | 0.337 |
| Eosinophils (%) | 1.77 | 1.28 | 1.81 | 1.20 | 0.0333 | -0.374–0.441 | 0.865 |
| Basophils (%) | 0.461 | 0.215 | 0.511 | 0.249 | 0.0500 | -0.0422–0.142 | 0.269 |
| Monocytes (%) | 5.00 | 1.10 | 5.09 | 1.10 | 0.0944 | -0.381–0.569 | 0.680 |
| Lymphocytes (%) | 33.6 | 6.67 | 34.7 | 7.12 | 1.11 | -1.30–3.53 | 0.345 |
| Blood biochemistry |  |  |  |  |  |  |  |
| AST (U/l) | 17.9 | 4.02 | 18.8 | 4.22 | 0.833 | -0.105–1.77 | 0.0782 |
| ALT (U/l) | 18.7 | 12.0 | 19.1 | 12.3 | 0.389 | -0.866–1.64 | 0.522 |
| γ-GT (U/l) | 35.9 | 49.2 | 34.4 | 45.8 | -1.50 | -3.93–0.932 | 0.210 |
| LDH (U/l) | 155 | 21.0 | 159 | 19.3 | 4.50 | -1.06–10.1 | 0.106 |
| Albumin (g/dl) | 4.76 | 0.248 | 4.71 | 0.271 | -0.0500 | -0.151–0.0513 | 0.312 |
| Urea nitrogen (mg/dl) | 13.4 | 3.50 | 12.2 | 2.46 | -1.18 | **-2.13 to -0.235** | **0.0174\*** |
| HDL cholesterol (mg/dl) | 68.5 | 11.8 | 68.3 | 9.97 | -0.222 | -2.69–2.25 | 0.852 |
| LDL cholesterol (mg/dl) | 114 | 40.2 | 112 | 36.3 | -1.72 | -6.12–2.68 | 0.420 |
| Triglycerides (mg/dl) | 92.2 | 61.2 | 95.6 | 74.5 | 3.39 | -35.8–42.6 | 0.857 |
| CRP (mg/dl) | 0.0489 | 0.0466 | 0.0606 | 0.0567 | 0.0117 | -0.00846–0.0318 | 0.238 |
| Cytokines |  |  |  |  |  |  |  |
| IFN-γ (pg/ml) | 3.51 | 2.87 | 4.51 | 2.94 | 1.00 | -0.337–2.34 | 0.133 |
| IL-6 (pg/ml) | 0.552 | 0.531 | 0.852 | 1.01 | 0.301 | -0.117–0.718 | 0.147 |
| TNF-α (pg/ml) | 1.82 | 0.851 | 2.36 | 1.33 | 0.539 | **0.183–0.896** | **0.00536\*\*** |
| IL-1β (pg/ml) | 0.173 | 0.574 | 0.558 | 0.924 | 0.386 | **0.0693–0.702** | **0.0198\*** |
| IL-18 (pg/ml) | 119 | 64.6 | 269 | 280 | 150 | **25.9–274** | **0.0207\*** |
| IL-2 (pg/ml) | 0.641 | 1.01 | 0.747 | 1.12 | 0.106 | -0.0217–0.234 | 0.0981 |
| IL-8 (pg/ml) | 291 | 685 | 663 | 933 | 372 | **159–585** | **0.00184\*\*** |
| IL-10 (pg/ml) | 0.255 | 0.203 | 0.298 | 0.199 | 0.0421 | -0.0635–0.148 | 0.412 |

MCV: mean corpuscular volume; MCH: mean corpuscular hemoglobin; MCHC: mean corpuscular hemoglobin concentration; AST: aspartate aminotransferase; ALT: alanine aminotransferase; γ-GT: γ-glutamyltransferase; LDH: lactate dehydrogenase; HDL: high-density lipoprotein; LDL: low-density lipoprotein; CRP: C-reactive protein; IFN: interferon; IL: interleukin; TNF-α: tumor necrosis factor-α; SD: standard deviation; CI: confidence interval. \* $P < 0.05$ , \*\* $P < 0.01$ , \*\*\* $P < 0.001$ .

**Figure 1.**
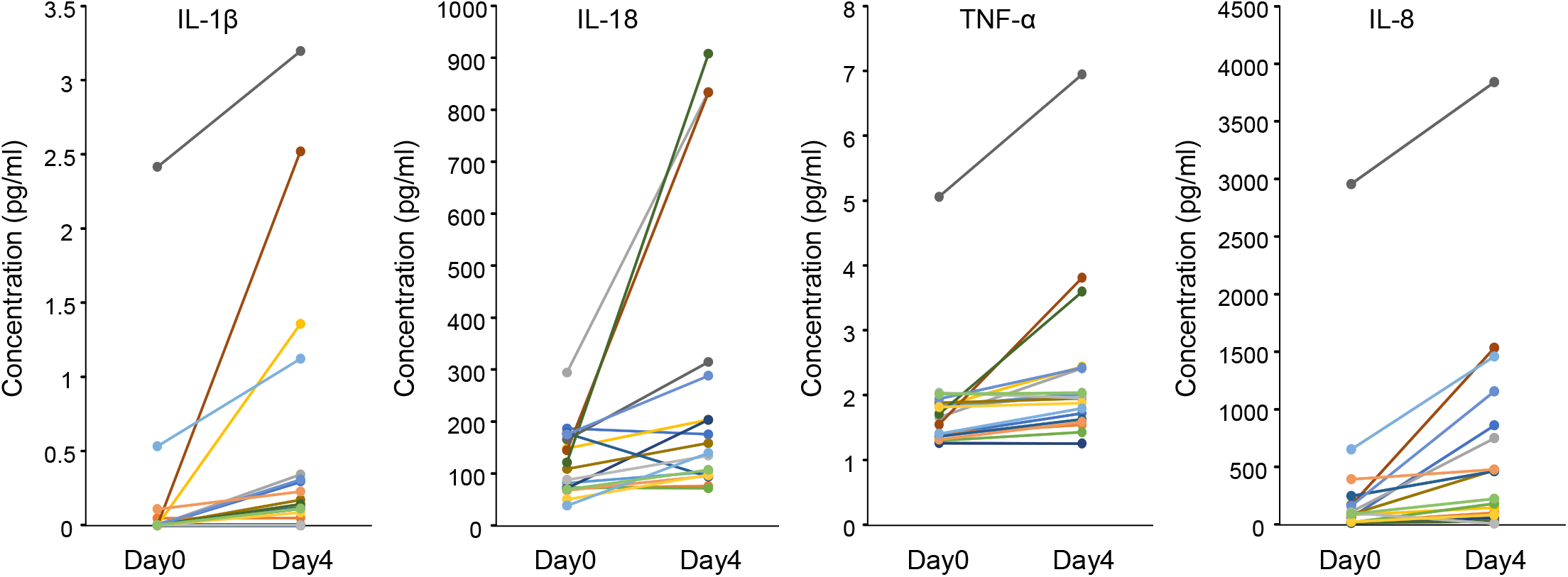
Changes in the plasma IL-1β, IL-18, TNF-α, and IL-8 levels before and after low-dose administration of QFPDT.

## Discussion

Inflammation is a host defense mechanism against invading pathogens. Acute inflammatory responses to the infection of single-stranded RNA (ssRNA) viruses such as SARS-CoV-2 are triggered by foreign ssRNA sensors such as Toll-like receptor (TLR) 7, TLR8 and NOD-like receptor, pyrin domain containing 3 (NLRP3) [7,8]. TLR7 and TLR8 are expressed mainly in dendritic cells and monocytes/macrophages and induce the expression of proinflammatory cytokines, including TNF-α and IL-6, in response to viral ssRNA [7]. NLRP3, when it recognizes viral ssRNA in the cytoplasm, initiates the inflammasome-mediated processing of pro-IL-1β and pro-IL-18 to release active IL-1β and IL-18 [7,9]. We speculate that QFPDT may create a blood cytokine conditions similar to those in acute antiviral immune responses.

TNF-α and IL-1β are well-recognized central regulators of acute inflammatory reactions. In particular, IL-1β activates neutrophil functions, such as migration, superoxide production, and survival. Conversely, neutrophils stimulates alveolar macrophages to release IL-1β during respiratory viral infection [10]. IL-18 also has diverse bioactivities; it activates both Th1–mediated cellular and Th2–mediated humoral immunity and enhances the cytotoxic activity of NK cells, which attack and eliminate virus-infected cells [9]. IL-8 is produced by a various types of cells such as monocytes/macrophages, fibroblasts and endothelial cells and serves as a potent chemotactic factor that induces the migration of neutrophils to the site of infection and enhances its phagocytic activity. In the respiratory system, IL-8 is produced by alveolar and airway epithelial cells, and IL-8–dependent inflammation in the lung is the first line of innate immune responses against respiratory pathogens [11].

In our trial, QFPDT had no significant effects on the plasma levels of IL-6 and IFN-γ. An excessive increase in IL-6 leads to severe immune dysregulation in COVID-19 patients [4–6]. IFN-γ is a critical mediator of Th1–dependent cellular immunity; however, a recent study has shown that IFN-γ stimulates the expression of the SARS-CoV-2 receptor ACE2 and thereby facilitates the viral entry into host airway epithelial cells [12]. The ineffectiveness of QFPDT on IL-6 and IFN-γ may thus be a preferable property for the prevention of CIVID-19.

In conclusion, our findings suggest that even a lower dose of QFPDT can upregulate a subset of proinflammatory cytokines without apparent adverse effects. However, since COVID-19 patients present with a broad range of inflammatory symptoms [4–6], this immunopharmacological property of QFPDT is apparently contradictory to its therapeutic benefits in the previous clinical trials [1–3]. We should therefore be careful in its prophylactic use in uninfected individuals until we have a better understanding of the pharmacological action of QFPDT through further clinical studies with larger cohorts.

## Data Availability

The data that support the findings of this study are available on request from the corresponding author, T.N. The data are not publicly available due to their containing information that could compromise the privacy of research participants.

## Funding

This research did not receive any specific grant from funding agencies in the public, commercial, or not-for-profit sectors.

## Disclosure of Potential Conflict of Interest

Y.K., K.A., K.K., M.M., and T.E. are employees of Takanawa Clinic. T.A. and T.N. have advisory roles in conducting clinical research in Takanawa Clinic and receive advisory fees from Takanawa Clinic.

## Author Contributions

Y.K., K.A., K.K., M.M., and T.E. contributed to the conception and design of the study, contributed to data acquisition, analysis, and interpretation, and critically revised the manuscript for important intellectual content. T.A. contributed to the conception and design of the study and critically revised the manuscript for important intellectual content. T.N. contributed to the conception and design of the study, contributed to data analysis and interpretation, and drafted the manuscript. All authors gave final approval of the submitted version of the manuscript and agree to be accountable for all aspects of the work.

## Notes

### Clinical Trial

UMIN000040341

### Author Declarations

All procedures were approved by the Ethics Committees of the Takanawa Clinic (approval number: 2020-2).

